# COVID-19 Neutralizing Antibody Surveillance Testing for Fully Vaccinated Individuals During Delta Variant Spread

**DOI:** 10.1101/2021.10.01.21264371

**Authors:** Jing Pan, Zhigang Li, Lin Wang, Joshua Szymanski, Maria Romano, Dylan Yin, Allen Wang, Thomas Small, Zhiying Zou, Jing Li, Greg Witham, Li Wang, Yubei Zhang, Kai Qi, Ray Yin

## Abstract

We recently performed 568 rapid neutralizing antibody (NAb) tests on 164 fully vaccinated individuals who received either Moderna or Pfizer COVID-19 vaccine regimens over 7 weeks. The NAb levels against the wild type (WA1/2020), Delta, and Kappa variants were measured and compared. Depending on each individual’s medical condition and vaccination status, the NAb levels for most of the fully vaccinated people decreased within 2-6 months, while a small number of individuals either generated non-detectable amount of NAbs after full vaccination (e.g., immunocompromised), or had high NAb levels lasting beyond 6 months. Since the NAb levels vary significantly among different individuals and decrease over time, the deployment of a low-cost rapid test to monitor NAb levels against both the wild type and emerging variants among fully vaccinated individuals can play a very crucial role to control the current pandemic. Our study provides an example of using such a rapid NAb test to fill this currently unmet medical need.

## Introduction

Due to the rapid spread of the SARS-CoV-2 Delta variant and increasing number of breakthrough cases reported^1,2,3^, the urgency to deploy a rapid and effective screening or surveillance test to identify those fully vaccinated individuals who are vulnerable to breakthrough infection is paramount. This group of people, once infected, can then transmit the virus to others including those who are fully vaccinated. Between June 29 and August 17, 2021, a surveillance testing study among fully vaccinated people with either Moderna or Pfizer vaccines was conducted using the NIDS^®^ COVID-19 Neutralizing Antibody (NAb) Rapid Test.

## Methods

The test is designed to detect neutralizing antibodies capable of blocking the binding between human Angiotensin-Converting Enzyme 2 (ACE2) and the SARS-CoV-2 virus spike Receptor Binding Domain (RBD) protein using a fingerstick blood sample. Results are interpreted within 15 minutes using a handheld reader. For this study, a competitive lateral flow immunoassay with a recombinant human ACE2 protein as a detector and an anti-RBD monoclonal antibody as a capture was used. The SARS-CoV-2 wild type (WA1/2020) spike RBD protein, as well as its variants including Delta RBD variant having L452R and T478K mutations and Kappa RBD variant with L452R and E484Q mutations were utilized to detect the neutralizing antibodies present in fingerstick blood samples collected from consenting, fully vaccinated or convalescent individuals. Due to the competitive assay format, the relative NAb level inversely correlates to the reader units obtained from the handheld reader. Specifically, the lower the reader unit value, the higher the NAb levels. Gene sequencing analysis was performed by Fulgent Genetics, Inc.

## Results

During this study, 568 NAb tests were performed on 164 fully vaccinated individuals with varying demographics as well as vaccination statuses over a 7-week period. The results showed that the NAb levels, although varied among different individuals, peaked around one to two week(s) after the second vaccine dose in both Moderna and Pfizer groups.

Even though the Limit of Detection (LOD) of the rapid test is ≤ 0.5 µg/mL based on the positive control antibody used for assay calibration, this concentration appears to be too low to be used as a cutoff point for the assessment/prediction of breakthrough infection risks. For example, at 1 µg/mL NAb level, three COVID-19 breakthrough cases between age 18 and 56 were identified and confirmed by both antigen and RT-PCR tests during this study. All such breakthrough individuals were within 2-4.5 months of their full vaccination. Gene sequencing analysis performed on two of the three samples with cycle threshold (Ct) values of 16.56 and 15.76, respectively, revealed that one was infected by the Delta (B.1.617.2) variant and the other by the Mu (B.1.621) variant. In terms of viral load over time, the individual infected by Mu was tested positive by antigen tests for 10 consecutive days, while the one infected by Delta was positive only for 3 consecutive days. All three breakthrough individuals had more than two COVID-19/flu like symptoms for 3-5 days, but no hospitalizations were required. The individual infected by Mu variant had a peak NAb level at about 2 µg/mL after full vaccination, which maintained at > 1 µg/mL for over 3 months. However, when tested 18 days before COVID-19 symptom onset, the NAb level decreased to between 0.5-1 µg/mL, as compared to the positive controls. To better assess the risk of breakthrough infection, the assay cutoff point was, therefore, set at 1.0 µg/mL or the High Positive Control (HPC) level. Using this cutoff, the negative-responder rate (NR%) based on the NAb test increased across both the Moderna and Pfizer groups, as well as from the wild type to Delta, and then to Kappa variants (**Table 1**). For the Delta variant, the NR% was 45% and 51% for the Moderna and Pfizer cohorts, respectively, while for the Kappa variant, it increased to 54% and 66%, respectively.

**Table 1.** Negative-responder rates for SARS-CoV-2 wild type (WA1/2020), Delta and Kappa variants, as tested by NIDS COVID-19 Neutralizing Antibody Rapid Test

| RBD Mutation | Moderna |  | Pfizer |  |
| --- | --- | --- | --- | --- |
|  | N | HPC | N | HPC |
| Wild Type (WA1/2020) | 93 | 32 (34%) | 122 | 52 (43%) |
| Delta | 86 | 39 (45%) | 107 | 55 (51%) |
| Kappa | 68 | 37 (54%) | 92 | 61 (66%) |

NAb titer measurement was further performed on some of the individuals with NAb levels above 1 µg/mL over time. It was found that the NAb titers for these participants decreased within 4 months from full vaccination with one individual decreased by as much as 8-fold during the same period (**Figure 1A**).

**Figure 1.**
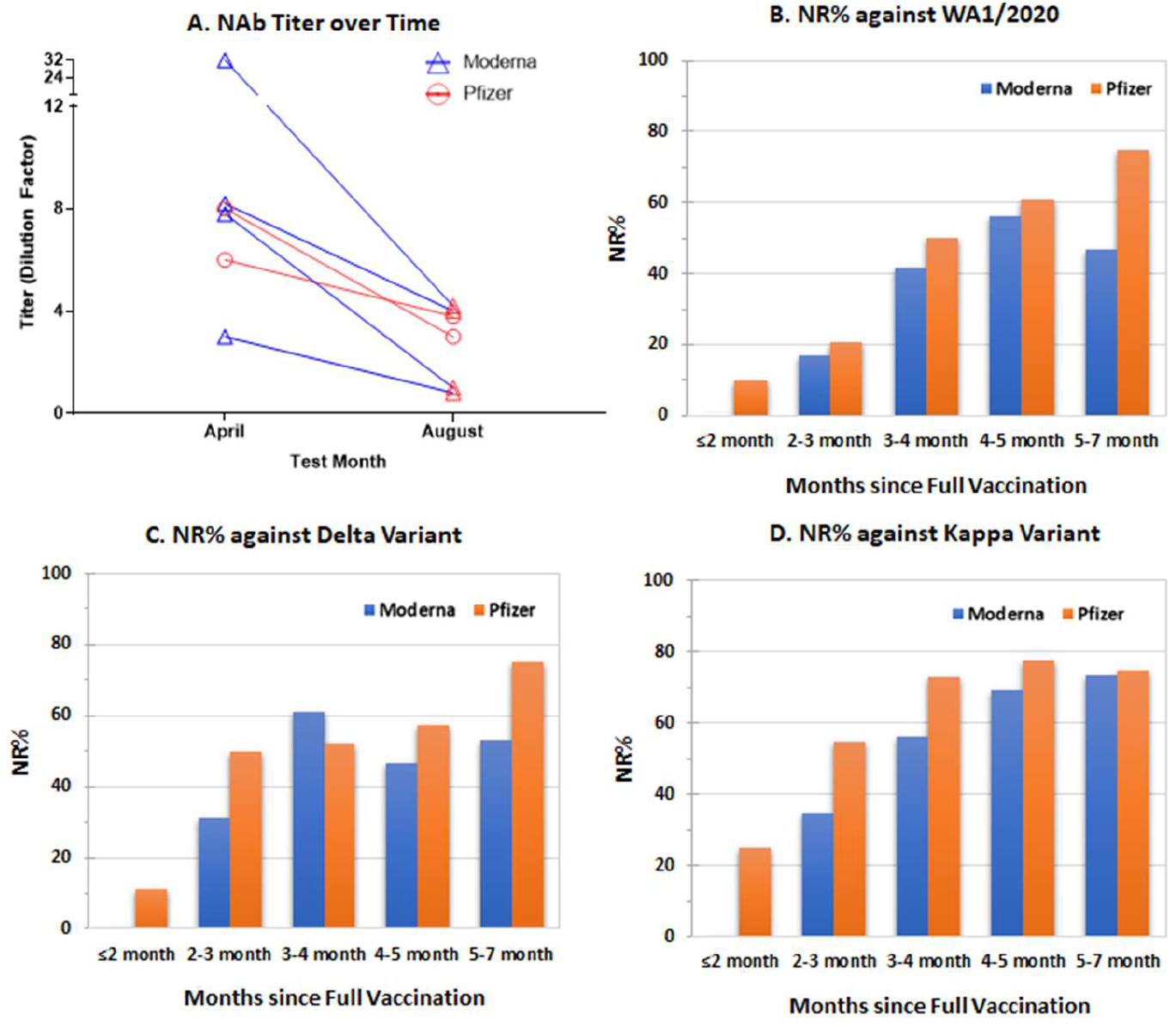
NAb test results among fully vaccinated individuals from both Moderna and Pfizer groups: **A**, NAb titer changes over time; **B**, Negative-Responder Rate (NR%) against Wild Type (WA1/2020) by vaccination status; **C**, NR% against Delta variant by vaccination status; and **D**, NR% against Kappa variant by vaccination status.

Longer vaccination time resulted in more negative-responders as compared to the positive control (**Figure 1B-1D**), while older age group also had higher numbers of negative-responders (**Figure S2**). Depending on each individual’s medical condition and vaccination status, the NAb levels for most of the fully vaccinated people decreased within 2-6 months, while a small number of individuals either generated non-deteactable amount of NAbs after full vaccination (e.g., immunocompromised), or had high NAb levels lasting beyond 6 months. For the wild type, it took 3-4 months after full vaccination for the NR% to reach to about 50% for the Pfizer group, while for Delta and Kappa, it only took about 2-3 months for the NR% to reach 50% for the same group. Overall, the Moderna group seemed to have slightly lower negative-responder rates over time, except for the 3-4 month group for the Delta variant, in which a higher NR% was observed in the Moderna group than Pfizer. This is because the former had 2 participants over 80 years old, while the latter had none.

## Discussion

The levels of NAb are highly predictive of immune protection for seven vaccines currently on the market^4^. The decrease in NAb levels among fully vaccinated individuals with longer vaccination time and/or older ages indicated that the vaccine-induced immunity is waning within 6 months, particularly for the Pfizer group. Depending on individual health condition(s), vaccination time, medications used, and age, some became negative-responders based on our assay cut-off in as early as 2 months, while others maintained their protection beyond 6 months. In addition, while the waning vaccine protection over time may have contributed to the higher breakthrough infection cases reported recently, it is interesting to note that some of these breakthroughs occurred within 2-4.5 months from full vaccination and were caused by Delta and Mu variants. This suggests that the presence of variants also played an important role in the recent surge of breakthrough infection cases among fully vaccinated individuals. These variations will make the decision on giving a booster shot to all fully vaccinated people at a fixed time point, e.g., after 6 months, more difficult. Therefore, the rapid NAb test can serve as a practical tool for monitoring NAb levels or vaccine effectiveness among fully vaccinated individuals during COVID-19 pandemic.

## Data Availability

Testing data are available from the corresponding authors upon request.

## Acknowledgements

The authors would like to thank Dr. Bing Zhang (MacroStat Inc), Dr. Qun Sun, Dr. Gann Xu, Ms. Renee Street, Ms. Wujun Li, Ms. Zhi Jiang, Mr. William Auerbacher, and Ms. Rosa Wong for their discussions and assistance. The development of NIDS^®^ COVID-19 Neutralizing Antibody Rapid Test is funded in part by US Army (special thanks to Ms. Janet Jensen for her guidance and support of this project). The authors would also like to thank Mr. Jason Cheng and Dr. Harry Gao (Fulgent Genetics, Inc.) for performing the RT-PCR and gene sequencing analysis for the vaccine breakthrough patient samples.

## Supplementary Appendix

### Testing Procedure

A lyophilized recombinant RBD recombinant protein, wild type, Delta or Kappa, was first reconstitute in an ANP assay buffer to generate a bulk testing solution. For each test, 100 µL of the reconstituted solution is mixed with a 20 µL of plasma or fingerstick whole blood sample. The resulting mixture is then incubated for 1 minute at room temperature, followed by dispensing the sample mixture to the test device sample well. After 15 minutes, the result was analyzed using a Stand-Alone-Reader 4 (SAR4) developed at ANP to provide a semi-quantitative result. Three positive controls at the Ultra Hight Positive Control (UHPC, 2.83 µg/mL), High Positive Control (HPC, 1 µg/mL), and Low Positive Control (LPC, 0.5 µg/mL) levels were used for initial analysis, and the final floating assay cutoff point was set at HPC (1 µg/mL) for the assessment of infection risks.

### NAb testing using known positive and negative samples

64 negative (Pre-Covid) and 30 positive (convalescent) human plasma samples were tested using the NIDS COVID-19 Neutralizing Antibody Rapid Test, along with 2 negative controls, 2 LPC, 2 HPC, and 2 UHPC positive controls, respectively. The randomized results are shown in **Figure S1** with 11/30 positive samples showing negative results on NIDS COVID-19 NAb test. All pre-COVID negative patient specimens showed consistent negative results on the NAb test with 0% false positives.

**Figure S1.**
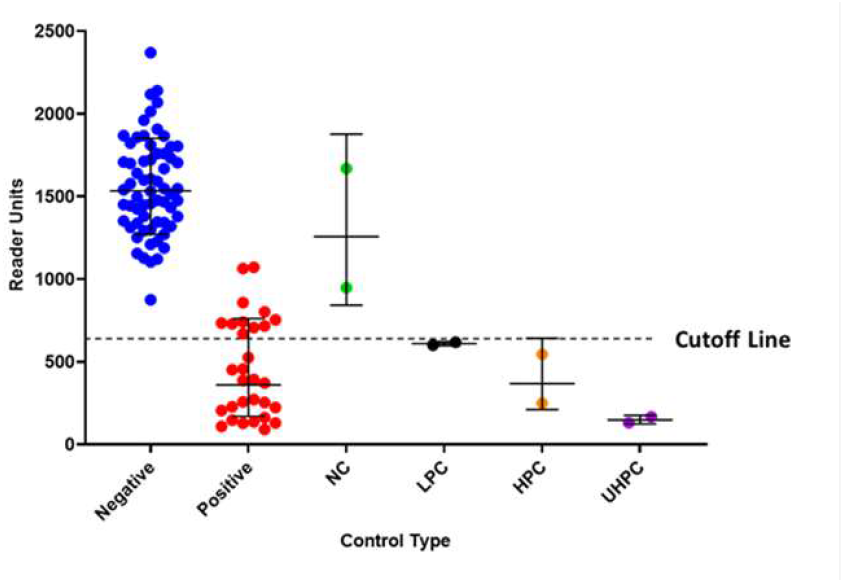
Test results of plasma samples (64 negative/pre-COVID-19 and 30 positive RT-PCR confirmed) as measured by NIDS COVID-19 neutralizing antibody rapid test. Negative: pre-COVID-19 plasma samples; Positive: convalescent plasma samples; NC: Negative Control; LPC: Low Positive Control; HPC: High Positive Control; UHPC: Ultra High Positive Control.

**Figure S2.**
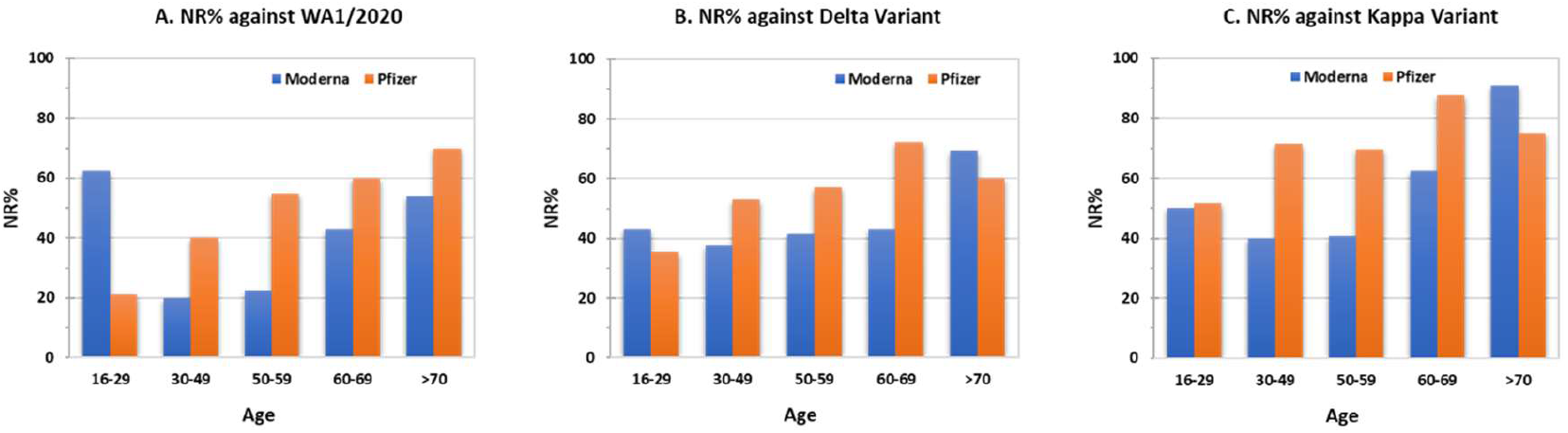
NAb test results among fully vaccinated individuals from both Moderna and Pfizer groups by age group: **A**, NR% against Wild Type (WA1/2020); B, NR% against Delta variant, and C, NR% against Kappa variant.

**Supplementary Table 1.** Demographics and characteristics of fully vaccinated Moderna and Pfizer-BioNTech participants from June 29 to August 17, 2021.

|  | Moderna | Pfizer |
| --- | --- | --- |
| **Demographics** |  |  |
| Number of people | 75 | 89 |
| Sex (female) | 41 | 46 |
| Sex (male) | 34 | 43 |
| Age (median, range) | 57 (21 - 86) | 49 (12 - 89) |
| Age group |  |  |
| 16-29 | 6 | 24 |
| 30-49 | 14 | 15 |
| 50-59 | 23 | 23 |
| 60-69 | 19 | 15 |
| >70 | 13 | 9 |
| **Vaccination status at the time of testing** |  |  |
| Days after the 1st dose | 145 (87 - 223) | 131 (31 - 217) |
| Days after the 2nd dose | 117 (59 - 195) | 110 (10 - 196) |
| **Race or ethnic group** |  |  |
| Asian (including Indian) | 35 | 42 |
| Black or African American | 5 | 9 |
| Hispanic | 0 | 3 |
| White or Caucasian | 35 | 35 |

